# Can the Rapid Antigen Test for COVID-19 Replace RT-PCR: A Meta-analysis of Test Agreement

**DOI:** 10.1101/2021.10.19.21265190

**Authors:** Ibrahim Elmakaty, Abdelrahman Elsayed, Rama Ghassan Hommos, Ruba Abdo, Amira Mohamed, Zahra Yousif, Maryam Fakhroo, Abdulrahman Alansari, Peter V. Coyle, Suhail A. R. Doi

## Abstract

**Background:** Several studies have compared the performance of reverse transcription-polymerase chain reaction (RT-PCR) and antigen rapid diagnostic tests (Ag-RDTs) as tools to diagnose SARS-CoV-2 disease (COVID-19). As the performance of Ag-RDT may vary among different products and viral load scenarios, the clinical utility of the Ag-RDT remains unclear. Our aim is to assess the diagnostic agreement between Ag-RDTs and RT-PCR in testing for COVID-19 across different products and cycle threshold (Ct) values.

**Methods:** An evidence synthesis and meta-analysis of Positive Percent Agreement (PPA) and Negative Percent Agreement (NPA) was conducted after an exhaustive search of five databases to locate published studies that compared Ag-RDT to RT-PCR and reported quantitative comparison results. After the screening, quality assessment, and data extraction, the synthesis of pooled estimates was carried out utilizing the quality-effects (QE) model and Freeman-Tukey double arcsine transformation (FTT) for variance stabilization. Subgroup analysis was also conducted to evaluate the tests’ diagnostic agreement across distinctive products and Ct-value thresholds.

**Findings:** A total of 420 studies were screened by title and abstract, of which 39 were eventually included in the analysis. The overall NPA was 99.4% (95%CI 98.8-99.8, I^2^=91.40%). The PPA was higher in lower Ct groups such as groups with Ct <20 and Ct <25, which had an overall PPA of 95.9% (95%CI 92.7-98.2, I^2^=0%) and 96.8% (95%CI 95.2-98.0, I^2^=50.1%) respectively. This is in contrast to groups with higher Ct values, which had relatively lower PPA. Panbio and Roche Ag-RDTs had the best consistent overall PPA across different Ct groups especially in groups with Ct <20 and Ct <25.

**Interpretation:** The findings of our meta-analysis support the use of Ag-RDTs in lieu of RT-PCR for decision making regarding COVID-19 control measures, since the enhanced capacity of RT-PCR to detect disease in those that are Ag-RDT negative will be unlikely to have much public health utility. This step will drastically reduce the cost and time in testing for COVID-19.

**Funding:** This research did not receive any specific funding.

## Introduction

Severe acute respiratory syndrome coronavirus 2 (SARS-CoV-2), which emerged as a novel human pathogen in China at the end of 2019, is responsible for coronavirus disease 2019 (COVID-19). It causes symptoms such as cough, fever, and severe pneumonia. The World Health Organisation (WHO) reported more than 200 million cases of COVID-19, including approximately 5 million deaths as of the 7^th^ of October 2021.^1^ The reference test for COVID-19 diagnosis is reverse transcription-polymerase chain reaction (RT-PCR) using nasopharyngeal or oropharyngeal swabs.^2^ It is designed to detect viral RNA and has high sensitivity and specificity.^3^ Contrarily, COVID-19 antigen-detection rapid diagnostic tests (Ag-RDTs) diagnose active infection by detecting SARS-CoV-2 viral proteins in upper respiratory swabs samples. Unlike RT-PCR, Antigen tests produce results in 15 to 30 minutes. In addition, they are cheaper, don’t require specialized laboratory personnel, and can be produced in larger quantities for large-scale deployment.^4^

Several studies have compared RT-PCR and Ag-RDTs as tools to diagnose COVID-19 infection. Studies differ in conclusions with some concluding that there is an overall excellent agreement between the two tests.^5,6^ In contrast, other studies suggest that Ag-RDT is not reliable enough to be used alone for COVID-19 diagnosis.^7^ In addition, the performance of Ag-RDT can vary among different products and viral loads. Different studies have stratified the patients into separate groups based on the Ct-values to try to account for viral load. Despite this, there has not been much consensus on the use of the Ag-RDT and thus this synthesis aims to evaluate Ag-RDTs diagnostic agreement across different products and viral load scenarios in comparison to RT-PCR, in order to make firm recommendations on its utility. This would be essential for enhancing control measures in the current pandemic.

There have been several previous attempts at evidence synthesis but, to our knowledge, all previous syntheses^8-12^ used diagnostic accuracy synthesis methods with RT-PCR designated as a gold standard and computed sensitivity, specificity, and other diagnostic indices. This may have lengthened this debate for two reasons: First, and most important, the synthesis method is completely different. Bivariate synthesis of sensitivity and specificity was used to account for their inverse correlation versus univariate synthesis of Positive Percent Agreement (PPA) and Negative Percent Agreement (NPA) in this paper. Second, the diagnostic accuracy perspective tends to then lead on to post-test probability concerns that are not really warranted and may make the interpretation of the applicability of Ag-RDTs difficult. Our meta-analysis is the first to assess the performance of Ag-RDT under the qualitative test framework suggested by the Clinical and Laboratory Standards Institute (CLSI).^13^

## Methods

### Search strategy and selection criteria

This was a rapidly performed evidence synthesis and meta-analysis; therefore, protocol registration was not deemed appropriate. An extensive search strategy was designed to retrieve all relevant articles published in PubMed up to 15 Mar 2021, using the following MeSH terms: “COVID-19” AND “Immunoassay” AND “Reverse Transcriptase Polymerase Chain Reaction”. To ensure comprehensiveness, other databases such as Embase, Google Scholar, EBSCO, and Scopus were similarly searched for additional relevant articles. No restrictions were imposed on searches, and duplicate articles were removed without the use of an automation tool. This study was reported in conformity with the Preferred Reporting Items for Systematic Reviews and Meta-Analyses (PRISMA) guidelines.^14^

The records that were obtained from the literature search were further considered through screening of titles and abstracts using the Rayyan platform.^15^ Two reviewers independently screened papers and any disagreements were resolved by consensus involving the whole team. The full texts of studies deemed potentially eligible were then retrieved and double-screened independently, with referral to a co-reviewer in case of a discrepant judgment.

We adopted broad eligibility criteria and included studies that used Ag-RDT as their index test, RT-PCR as their reference test to detect COVID-19, and reported the values of Ag-RDT positives among RT-PCR positives, and Ag-RDT negatives among RT-PCR negatives. Studies that did not meet these inclusion criteria were excluded. We excluded studies where PPA and NPA could not be computed as well as literature reviews, recommendation studies, and studies not available in English.

### Data analysis

We extracted data from each eligible study on study design, study population, swab source, reference test, index test, and the number of positive Ag-RDT tests among positive RT-PCR and negative Ag-RDT tests among negative RT-PCR. All data were extracted into an excel worksheet (supplementary).

The Quality Assessment Tool for Diagnostic Accuracy Studies (QUADAS-2)^16^ was used to assess the methodological quality of selected articles, which was also done in an independent manner by two reviewers with the consultation of a third reviewer if needed. Even though this tool may not apply directly to diagnostic agreement studies, it was the closest in context and still provided an indication of the relative ranking of studies in terms of their methodological rigour. We deemed the relevant questions in the tool to refer to safeguards against bias, then counted the number of safeguards implemented in each study rather than assessing the risk of bias as a judgment as recommended by the tool.^16^ Thus, a numeric value of 1 was given if implemented (“yes/low bias”), and 0 if not implemented (“no/high bias/unclear”). This was done to allow the quality assessment to have quantitative utility^17^ for input into a bias-adjusted meta-analysis model.^18^ Besides excluding all judgment questions, we also excluded the following safeguard: “Is the reference standard likely to correctly classify the target condition?” as it did not apply to this agreement study.

The quality-effects (QE) model^19,20^ was utilized for bias-adjusted synthesis using the results of the quality assessment as relative ranks.^21^ The QE model accounts for methodological quality-related heterogeneity by redistributing study weights by quality rank, thus bias adjusting the synthesis.^19,20^ Pooled estimated PPA and NPA along with their 95% confidence intervals (95%CI) were calculated after utilizing the Freeman-Tukey double arcsine transformation (FTT)^22^ to stabilize the variance since double arcsine transformation has better performance in the synthesis of proportions.^23^ All results were back-transformed to proportions and converted to percentages for reporting. Heterogeneity was evaluated using the I^2^ statistic,^24^ while publication bias and possible small study effects were assessed by Doi plots, and the Doi plot symmetry was quantified using the LFK index.^25^ All analysis, graphs, and plots were undertaken using Stata version 16^26^ after installing the *metan* module^27^ as recently recommended.^28^

To compare Ag-RDT results across products and distinctive Ct-value thresholds, five groups were created according to Ct-values ranges. The first three groups are comprehensive ranges that altogether contain all Ct-values. Group one is Ct <20, group two is any range from 20 to 30 cycles, while group three is Ct >30. The fourth group included data restricted to Ct <25, while group five was restricted to Ct <30 (table 1). Ag-RDTs were also classified into separate groups by the product to identify which one had the best performance. A product was analysed individually only if it had five data sets or more by Ct-value group otherwise it was added into a combined products’ group called “other” (supplementary).

**Table 1:** Ct value range per group.

| **Ct group** | **Ct range** |
| --- | --- |
| Group 1 | Ct <20 |
| Group 2 | Ct 20-30 |
| Group 3 | Ct >30 |
| Group 4 | Ct <25 |
| Group 5 | Ct <30 |

## Results

The selection process of studies is depicted in a PRISMA flowchart (figure 1).^14^ Our search strategy yielded 416 studies and simultaneous search on other databases yielded 61 additional studies. 57 duplicates were identified and removed leaving a total of 420 studies that had undergone screening by title and abstract. Of which, 65 studies were assessed further for eligibility using their full-text articles. Only 39 studies were finally selected for inclusion in the analysis. The rest of the studies were excluded for reasons that are shown in the PRISMA chart.

**Figure 1:**
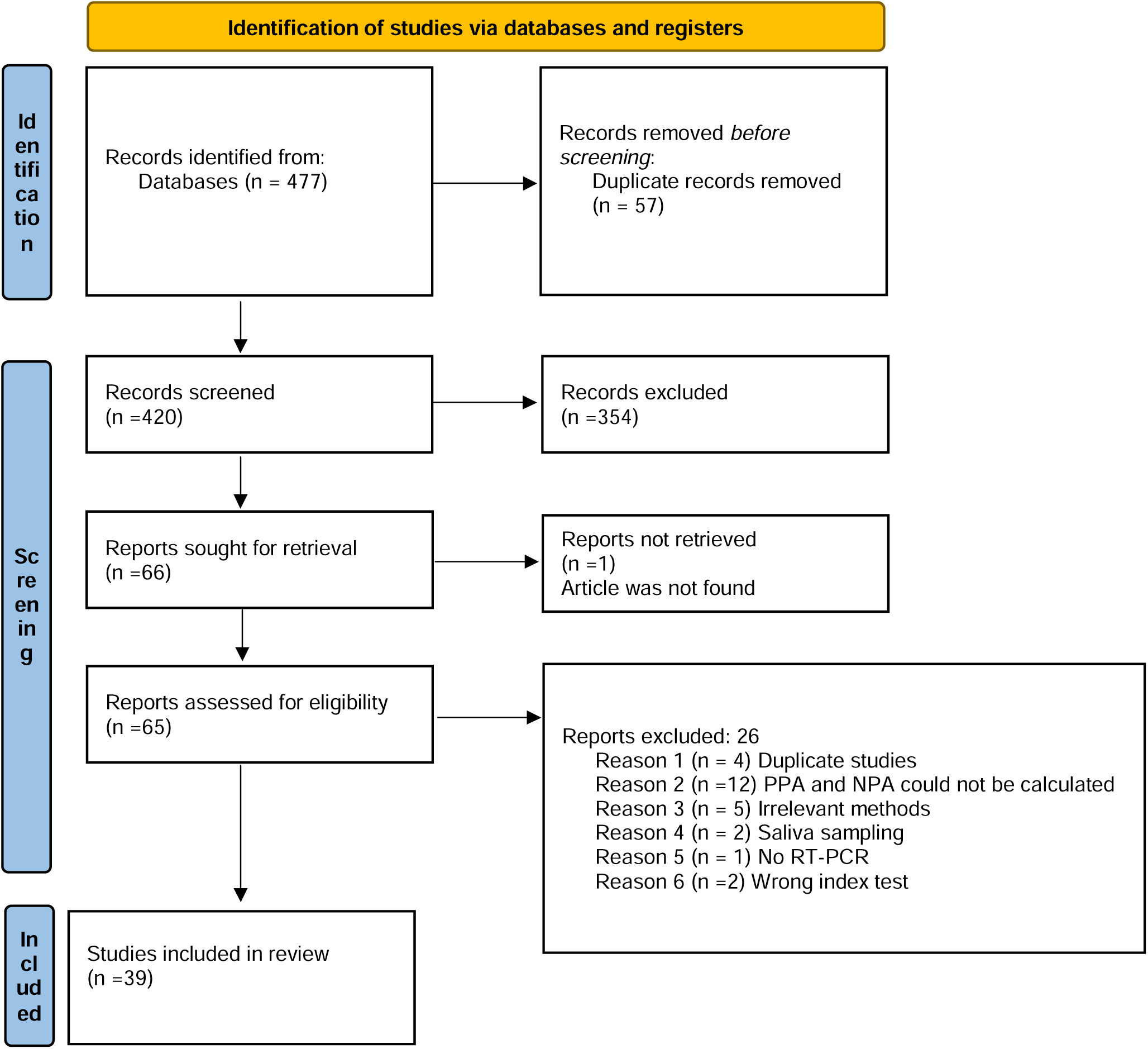
PRISMA flow-chart.

The characteristics of included studies are presented in table 2. The included studies came from various countries with the greatest number of studies from the USA (n=8), followed by Spain (n=6), and Japan (n=6). Participants in the included studies varied from being either symptomatic only (n=9), asymptomatic only (n=2), or a mix of both (n=25). The rest of the studies (n=3) and a subgroup in one of the studies^61^ did not provide information regarding the participant’s symptom status. One study was performed among pediatric patients only,^44^ while another study had a subgroup of pediatric patients that were analysed separately.^38^ One study had no information regarding the participants’ ages.^66^ The rest of the studies (n=36) had either adults only or participants of all ages. Three studies are reported as a and b (supplementary) based on either the participants’ age,^38^ methodology of participants selection,^61^ or the site where the sample collection took place.^63^ Some studies (n=4) have tested more than one Ag-RDT on the same cohort of patients, two studies had four Ag-RDTs,^53,59^ one had three Ag-RDTs,^51^ and two had two Ag-RDTs.^41,62^

**Table 2:**
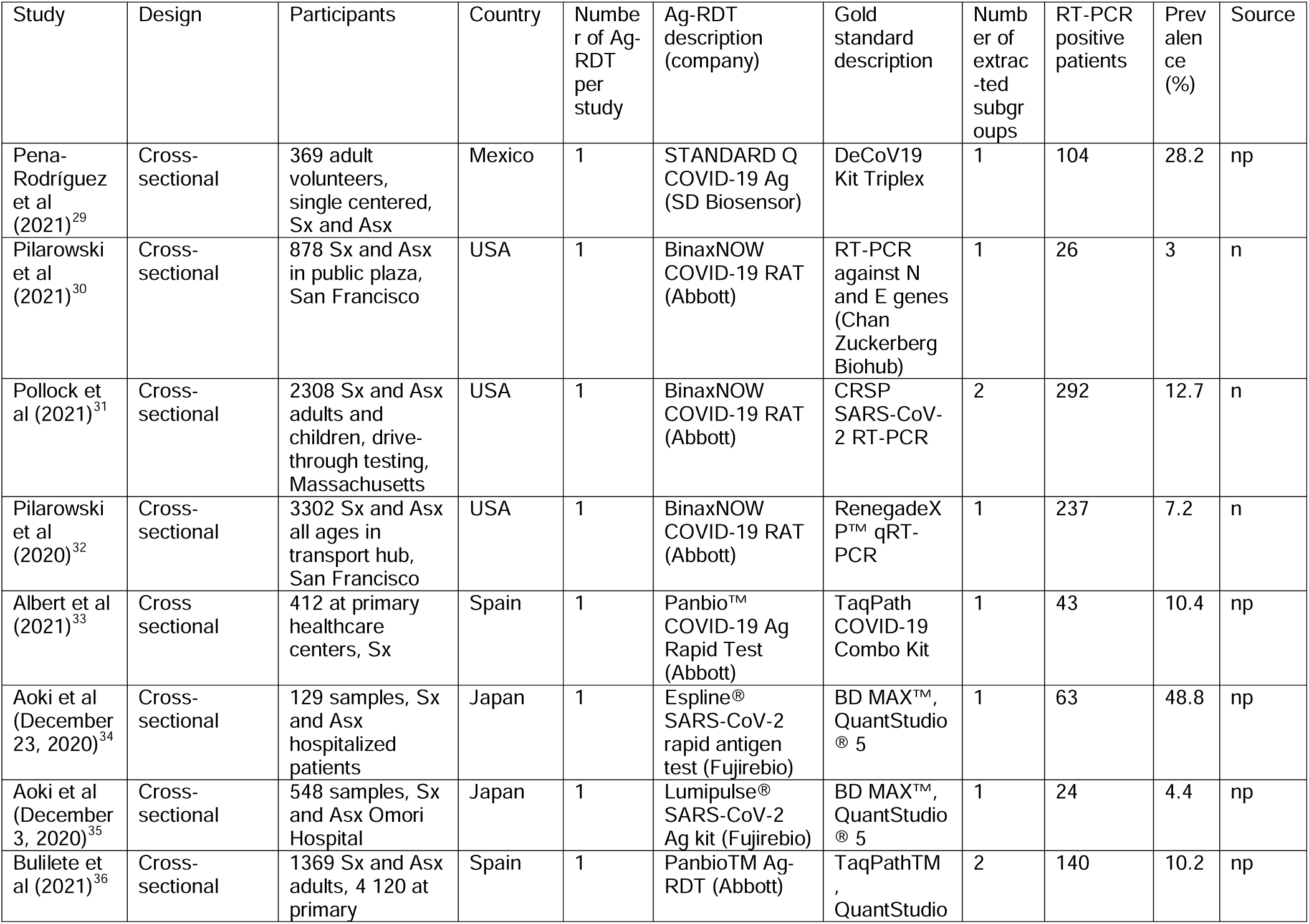
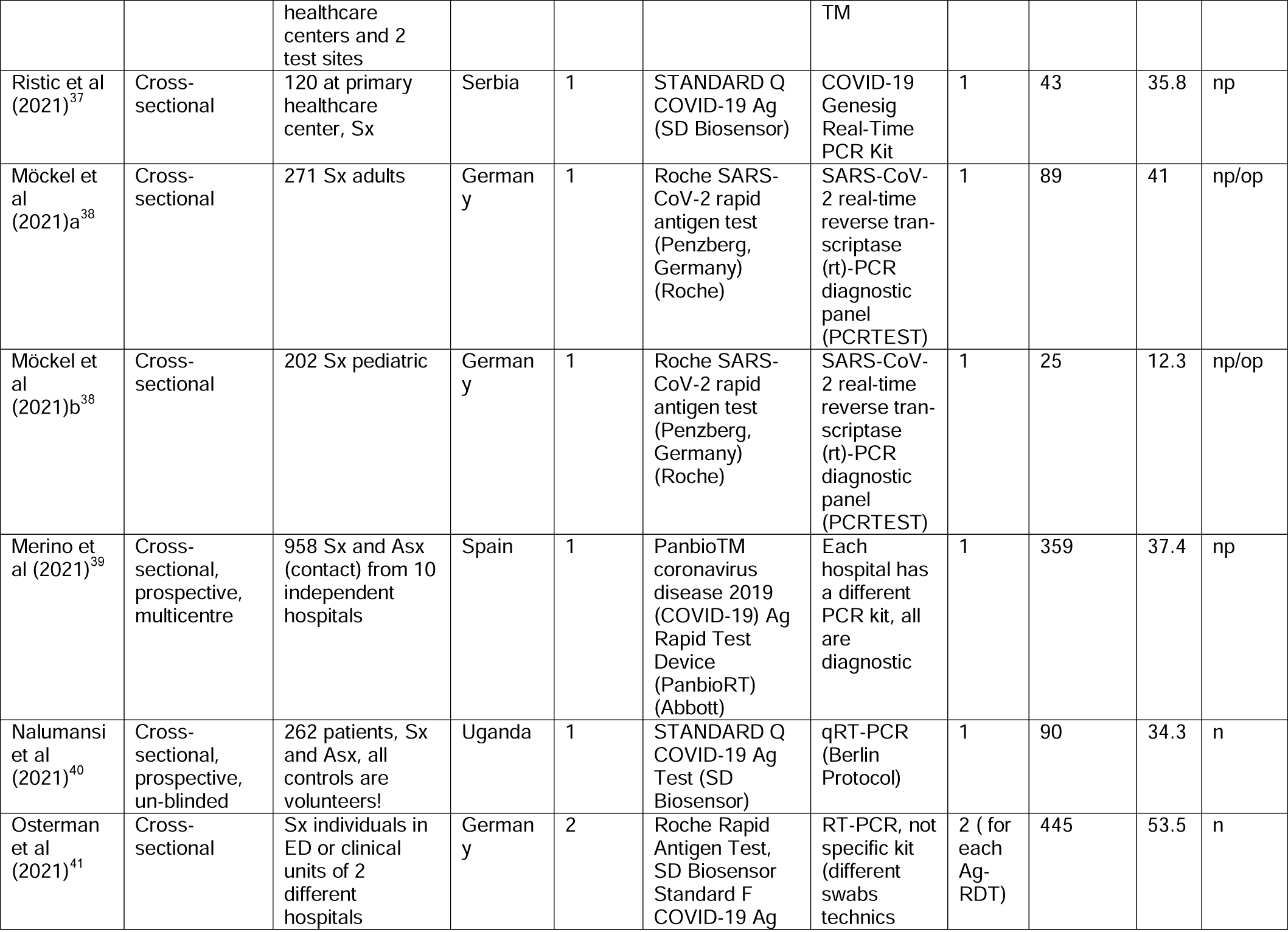
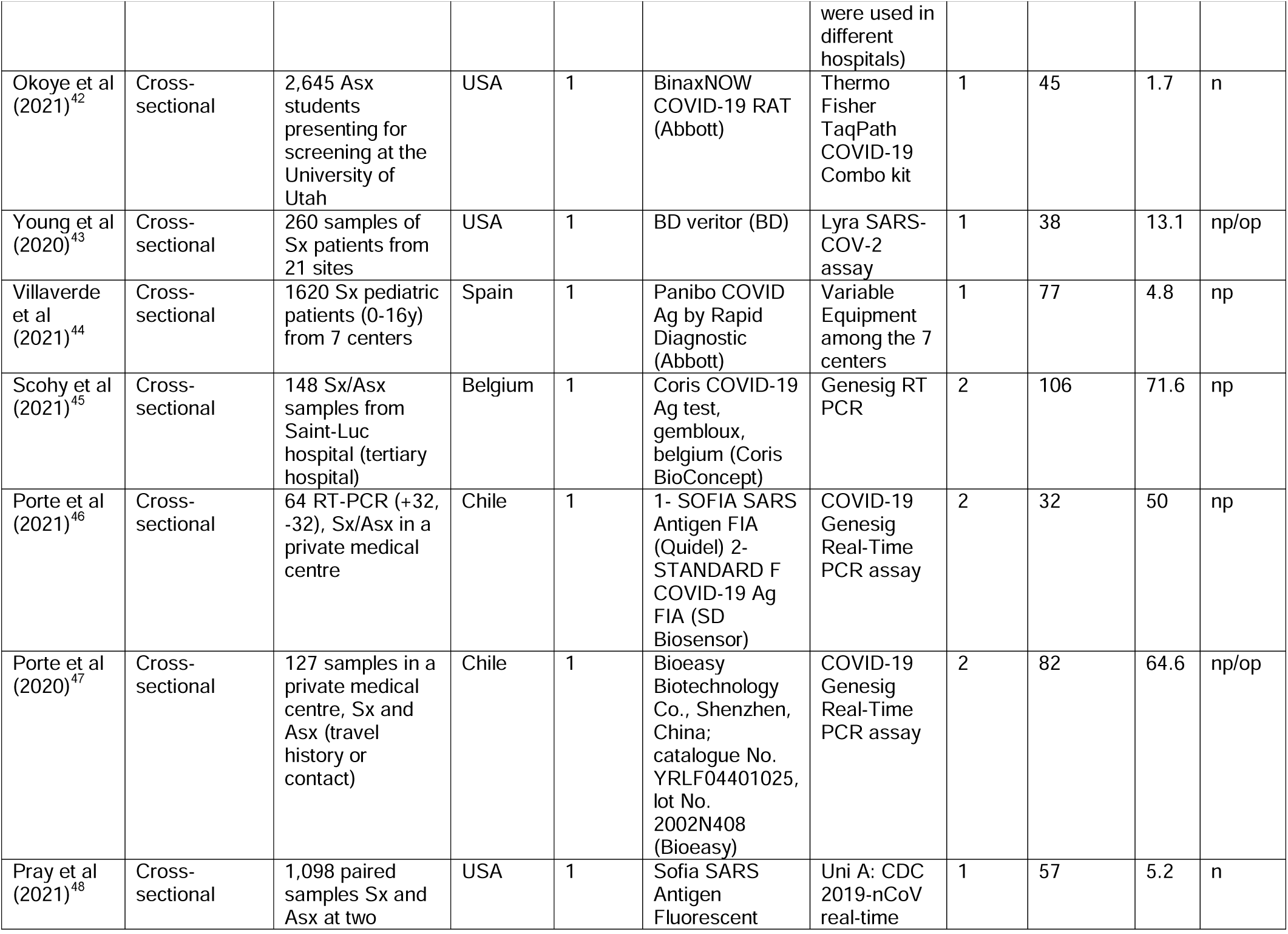
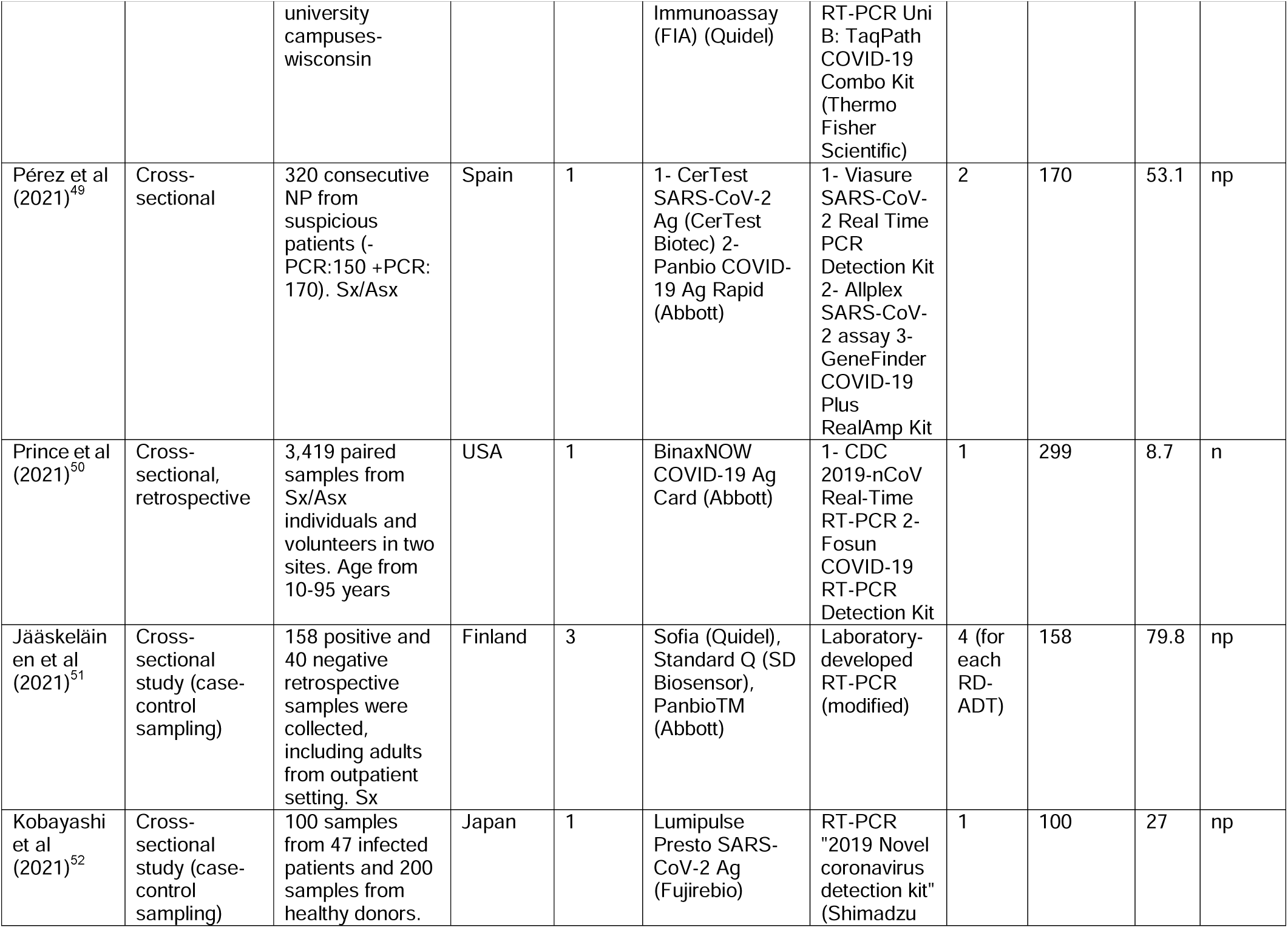
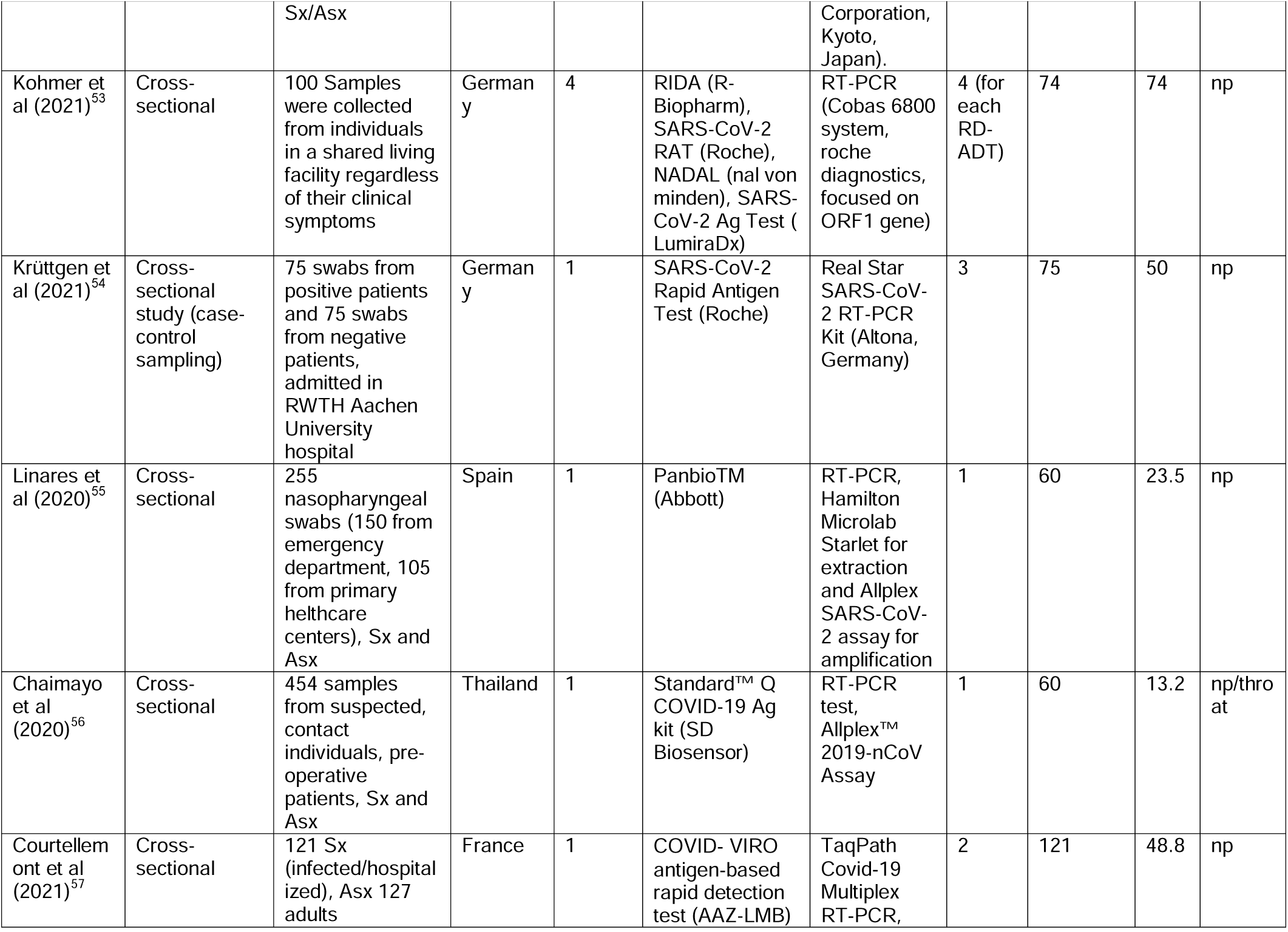
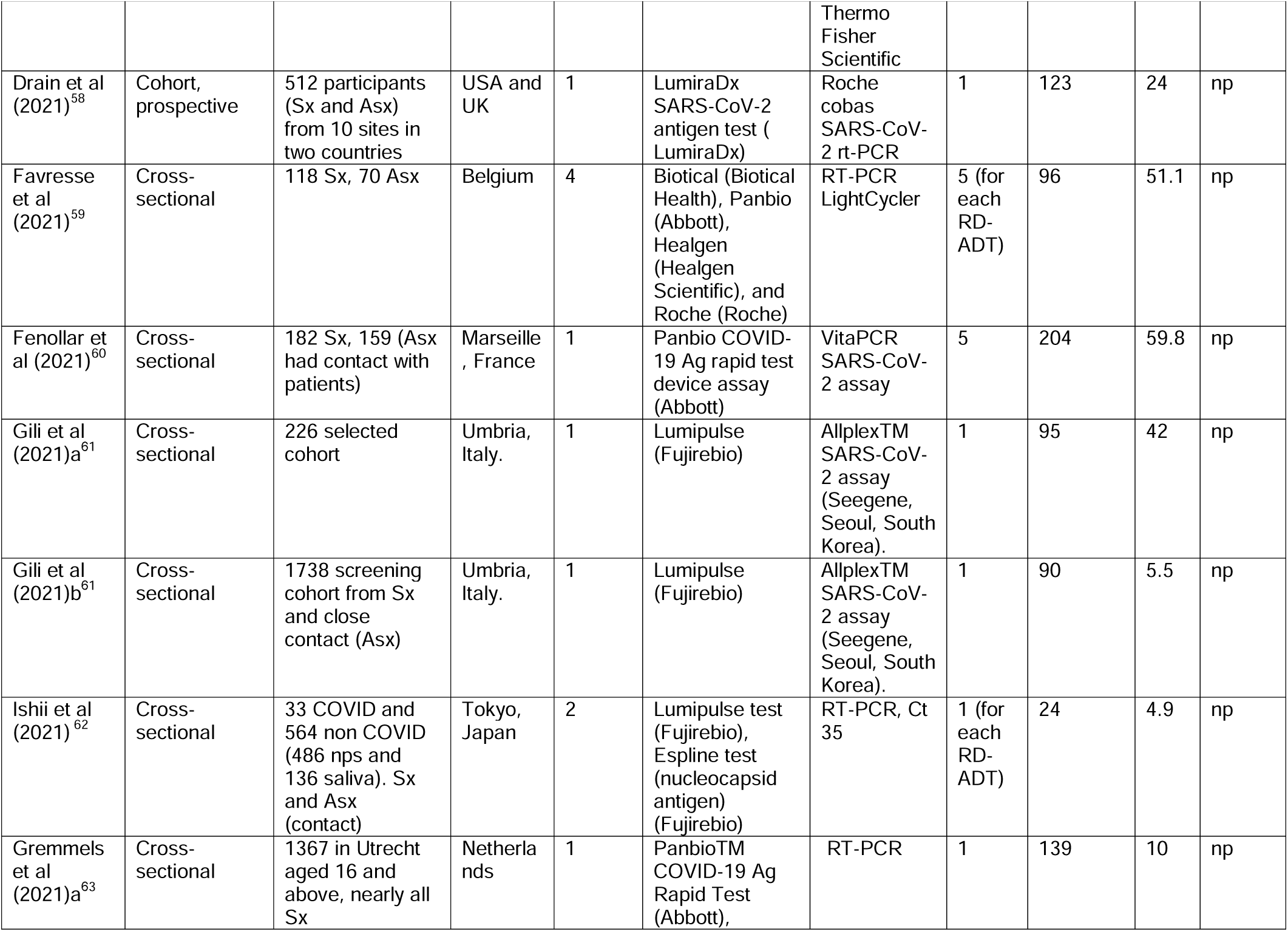
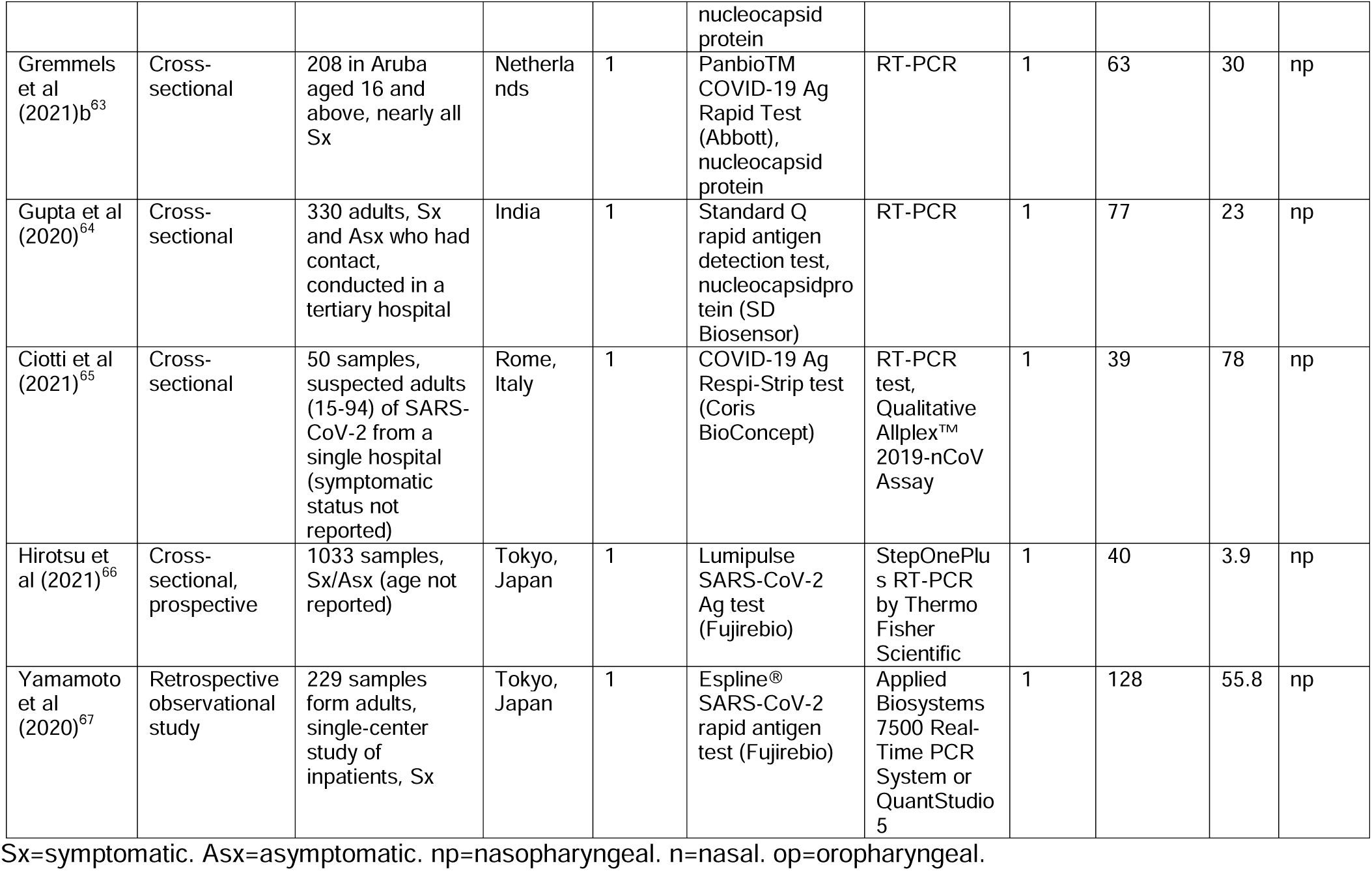
characteristics of included studies.

### Meta-analysis results

Since NPA is an estimate of COVID-19 negative population and is not expected to change across different Ct-value ranges, therefore only NPA at detection threshold is considered. Figure 2 summarizes the NPA results of different Ag-RDTs across all products. The analysis showed that the overall NPA was 99.4% (95%CI 98.8-99.8, I^2^=91.40%). There was negative asymmetry on the Doi plot (LFK index=-3.81; supplementary) suggesting that more of the smaller studies had a lower NPA. Despite the latter, NPA ranged from 99.9% (95%CI 99.7-100) to 97.9% (95%CI 91.9-100) for BinaxNOW and Lumipulse respectively, indicating high and consistent NPA values across products despite a downward bias from small studies.

**Figure :**
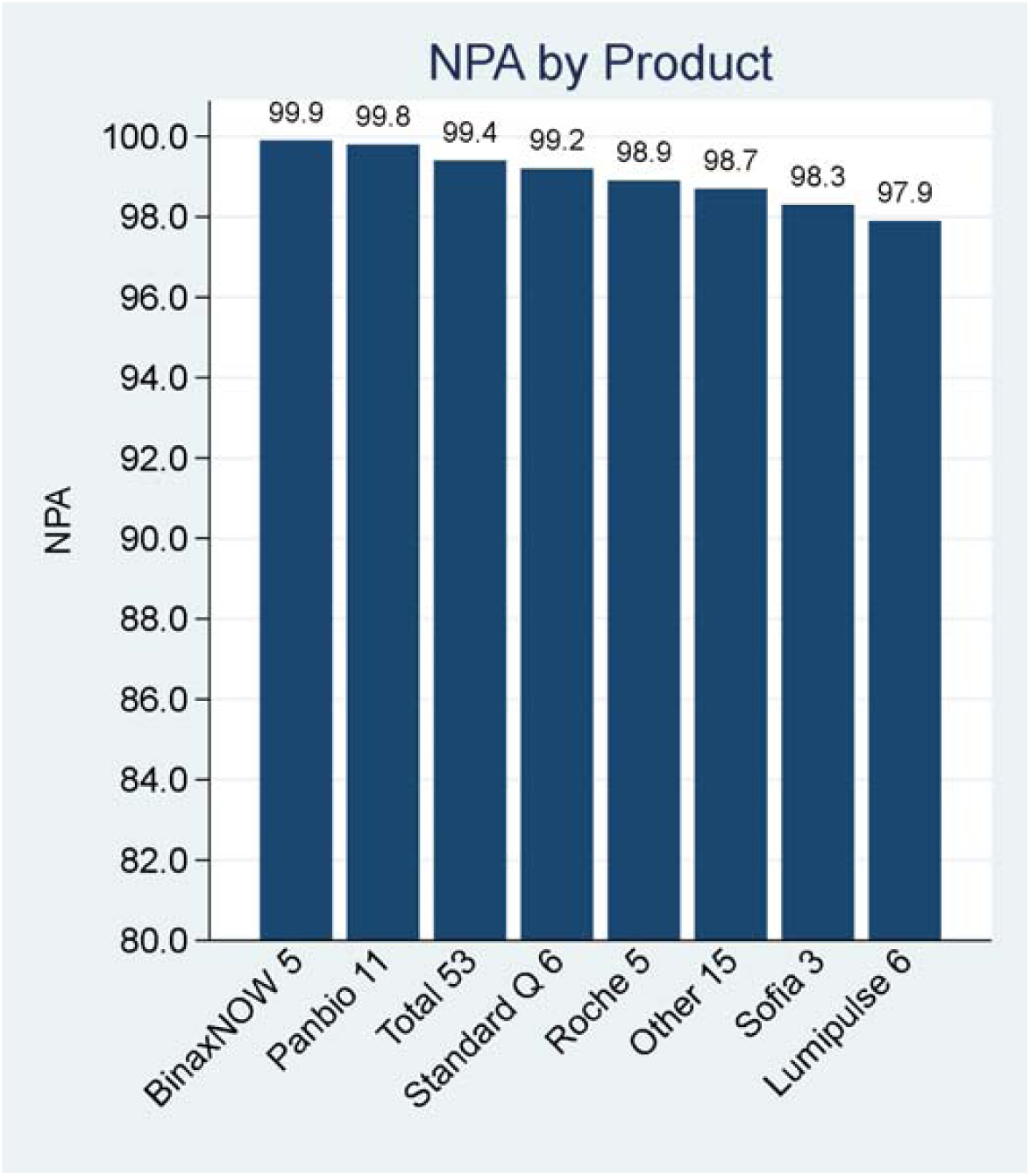
NPA for Ag-RDTs.

Figure 3 summarizes the PPA findings. The analysis showed better PPA values in groups with lower Ct-values (higher viral load). The overall PPA in the group with Ct <20 was 95.9% (95%CI 92.7-98.2, I^2^=0%) and the Doi plot suggested minor negative asymmetry (LFK= -1.29; supplementary). Similarly, the overall PPA of the group with Ct <25 was 96.8% (95%CI 95.2-98.0, I^2^=50.1%; figure 4) with minor negative asymmetry (LFK=-1.73; supplementary). As the Ct-value increases in groups with Ct <30 and Ct >30, the PPA drops significantly to 77.1% (95% CI 69.5-83.9, I^2^= 95.9%) and 38.6% (95% CI 12.9-67.7, I^2^=95.9%), respectively. There was only major negative asymmetry for Ct >30 group suggesting that some small studies had extremely low PPA values.

**Figure :**
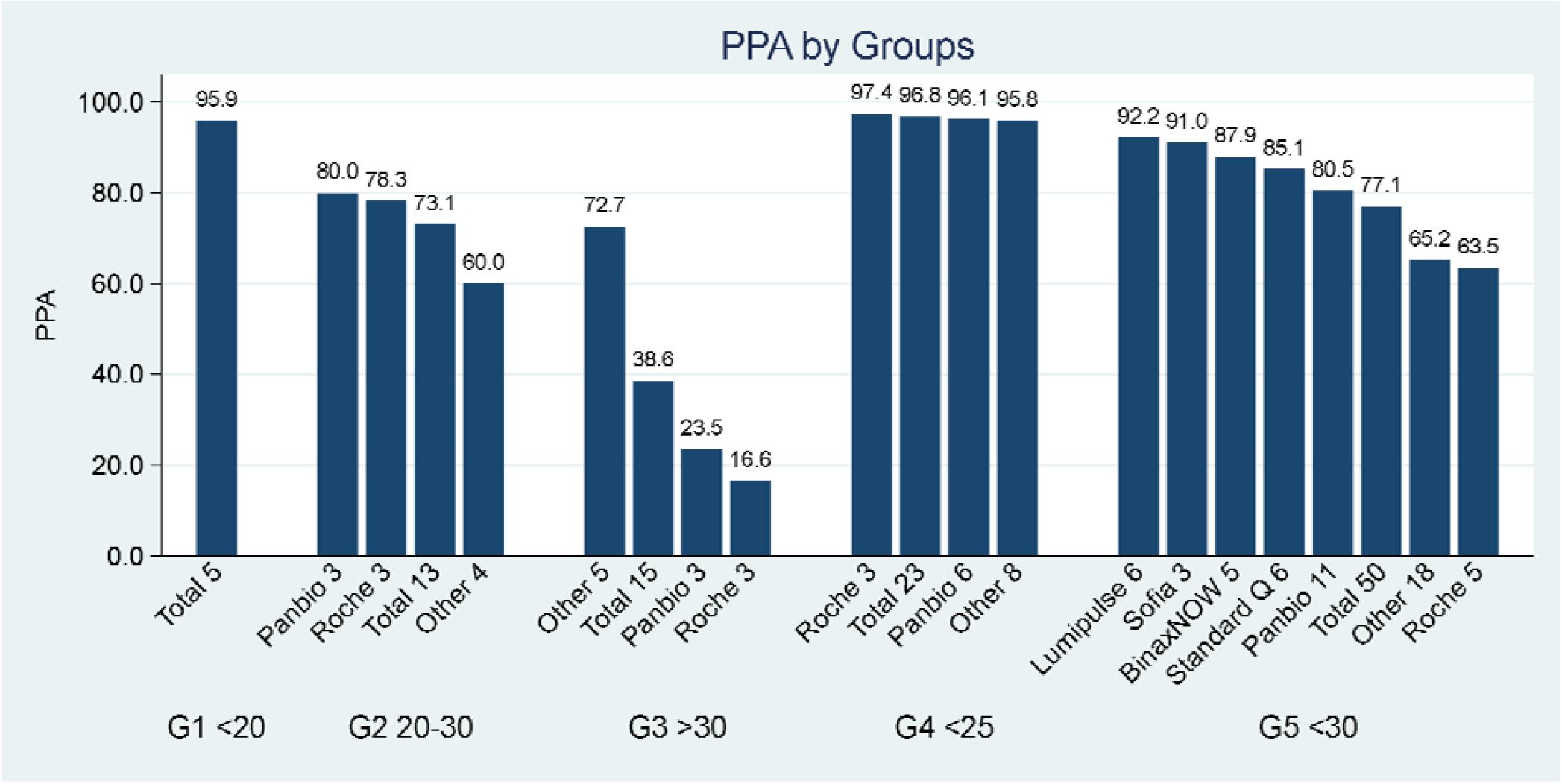
PPA for Ag-RDTs across Ct groups.

**Figure 4:**
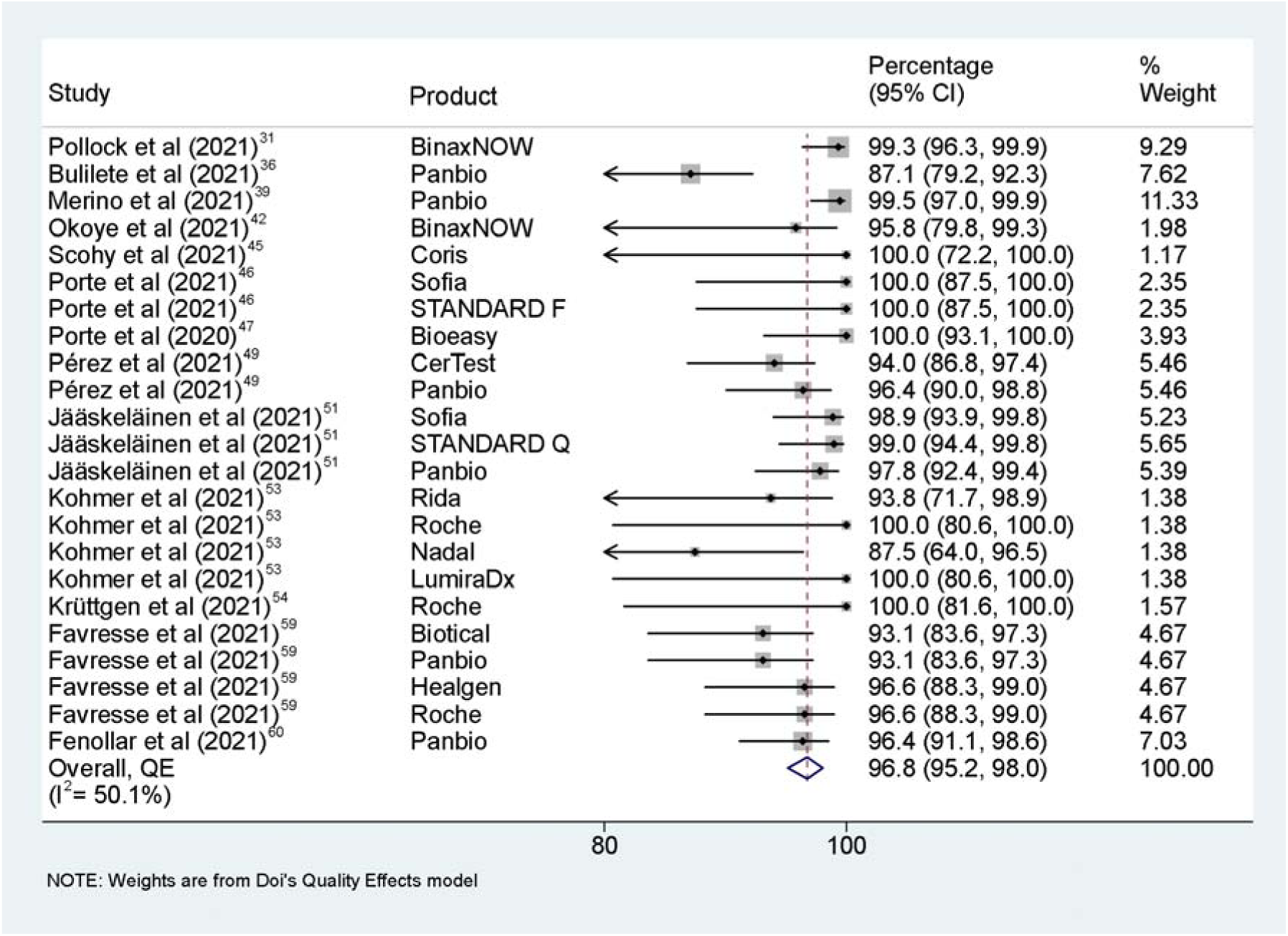
Forest plot for PPA of studies in group 4 Ct <25.

Regarding the PPA of particular products, the PPA of Panbio and Roche Ag-RDTs were the most consistent across clinically important groups such as group one and four (see figure 3).

## Discussion

The clear presence of an inverse relationship between the Ct value of a patient group and the PPA of Ag-RDT was demonstrated by our results (figure 5). What has not till now been clarified is where the PPA threshold lies, and we show this to be at Ct <25 with Panbio and Roche being the products demonstrating the highest PPA of 96.1% (95%CI 91.9-98.9) and 97.4% (95%CI 92.9-99.8), respectively.

**Figure 5:**
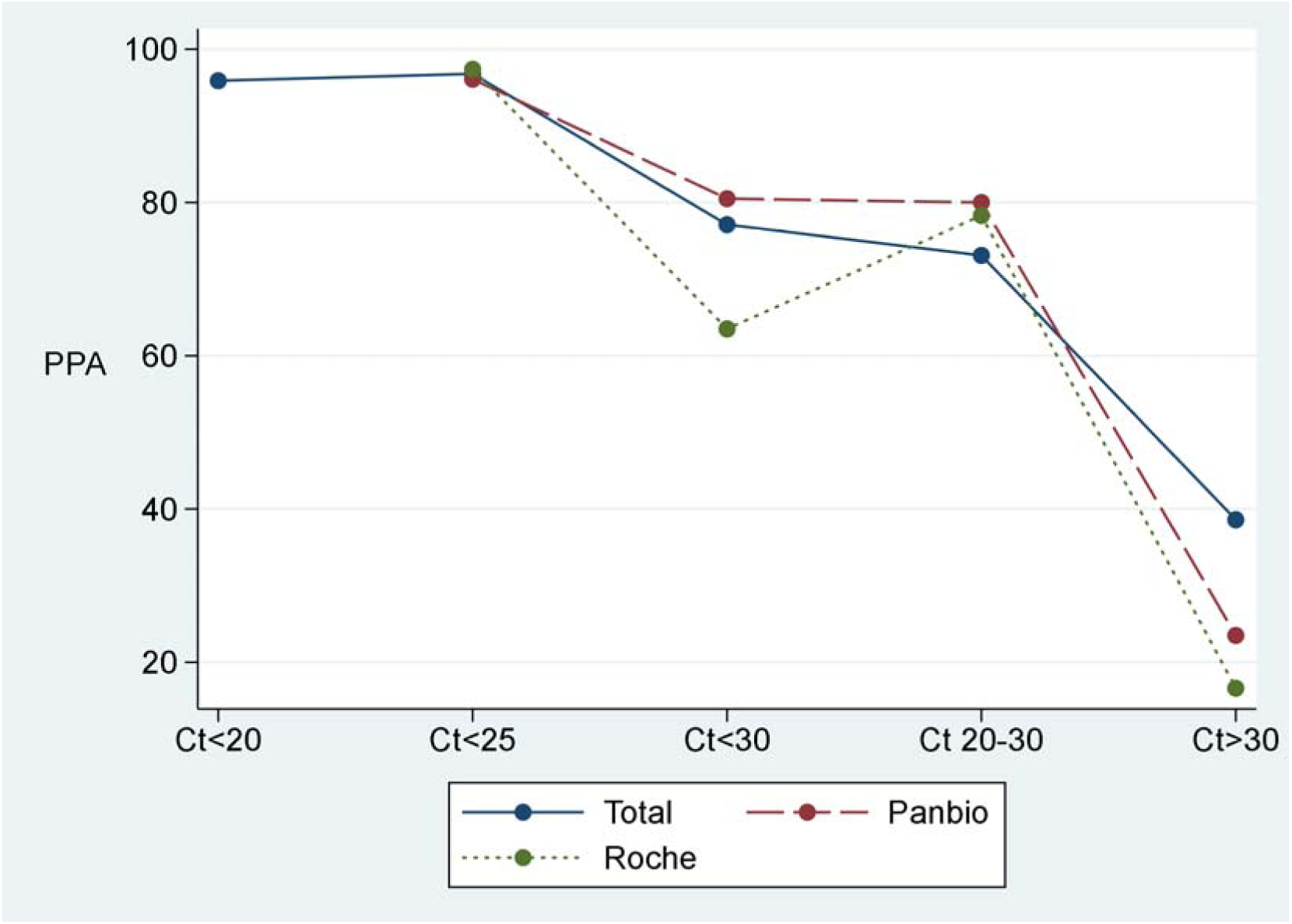
Line plot for overall PPA across Ct groups.

Beyond a Ct value of 25, the concordance of a positive Ag-RDT with RT-PCR declines. This drop-off may actually be an advantage rather than a limitation of the Ag-RDT since it has been reported that high Ct-value has a reduced chance of successful culture and of being contagious,^68^ which has been shown to be between Ct ranging from 24–30.^69,70^ This suggests that individuals start being more contagious at the threshold where the Ag-RDT operates best. Therefore, in case an Ag-RDT fails to detect an infected individual of Ct >25, it is likely that the individual is either noncontagious or is in early infection, which can be resolved by re-testing on subsequent days for contacts of a documented case.

In our study, we considered the RT-PCR to be a reference test, rather than a gold standard as done by previous meta-analyses targeting the same topic. This makes our study the first meta-analysis to have presented the PPA and NPA of Ag-RDTs, which is a primary strength of this study and allows interpretation of results for decision-makers. In addition, we used robust methodologies^19^ and broad yet specific selection criteria. It is also important to note that although this is an agreement study, we have found QUADAS-2^16^ to be suitable for quality assessment of diagnostic agreement studies with only the exclusion of a single safeguard.

There are a few limitations to our meta-analysis that should be mentioned. To begin with, not enough studies have stratified their analysis based on Ct-value, which resulted in data limitation clearly shown in our results. An example would be the low number of studies per product in group 1 (Ct <20), in which case we did not stratify by product in that group. Furthermore, most of the studies were earlier studies that evaluated patients with the classic variant of SARS-CoV-2 and thus tests should be selected based on the manufacturer having approved these for use with the newer variants of the virus.

The results of this study have several implications for practice. First, we recommend the use of Ag-RDT for trace and test procedures in the current pandemic thus replacing RT-PCR. Second, we suggest prioritizing Panbio™ COVID-19 Ag Rapid Test or Roche SARS-CoV-2 Rapid Antigen Test until further data is available or where local validation supports alternatives. Third, each health system must crosscheck results with variants under circulation and must make sure that the manufacturer actually has considered this in the product development and its futureproofing as is the norm for RT-PCR assays. This change is expected to result in huge cost and time savings for health systems worldwide and avoid quarantine of individuals who are unlikely to transmit the disease.^70^ We also recommend the use of Ag-RDT in asymptomatic patients in the first 5 days of contact as well as for screening purposes in public settings, such as sports matches or airports. This recommendation is in conformity with multiple studies showing high Ag-RDT performance in asymptomatic individuals with low viral loads.^30,32,36^ Finally, we suggest that manufacturers consider scaling up the use of Ag-RDTs by making them applicable to batch in large numbers, then have their results electronically transmitted to hospital information systems. This does not mean that point of care testing should be abandoned, but rather that there be a choice depending on what use is to be made of the test.

## Data Availability

The data used for analysis in this study is available from the corresponding author on reasonable request.

## Contributors

IE*, AE*, RGH*, RA*, AM, ZY, MF, and SAD provided critical conceptual input. SAD designed the methodology. IE*, AE*, and SAD performed the formal analysis. IE*, AE*, RGH*, RA*, PVC, and SAD validated the results of the study. IE*, AE*, RGH*, RA*, AM, ZY, MF, and AA were involved in the investigation, data curation, and writing the original draft. IE*, AE*, RGH*, RA*, AM, ZY, MF, PVC, and SAD reviewed and edited the report. RGH* was the administrator, and supervised this work along with SAD and PVC.

*Contributed equally to this report.

