## Supplementary Material for "Can the Rapid Antigen Test for COVID-19 Replace RT-PCR: A Meta-analysis of Test Agreement"

**Table of Content:**

**Tables	4**

**Doi plots	12**

**References	15**

**Additional information**

**Preliminary systematic search for finding relevant articles:**

Concept 1: COVID-19

Keywords:

"COVID-19"[Mesh] OR “COVID 19”[tiab] OR “Coronavirus Disease 2019”[tiab] OR “Coronavirus Disease-19”[tiab] OR “Coronavirus Disease 19”[tiab] OR “SARS-CoV-2 Infection*”[tiab] OR “2019-nCoV Infection*”[tiab] OR “2019 nCoV infection*”[tiab] OR “SARS Coronavirus 2 Infection*”[tiab] OR “2019 Novel Coronavirus Disease” [tiab] OR “2019 Novel Coronavirus Infection*”[tiab]

MeSH: "COVID-19"[Mesh]

Concept 2: Rapid antigen test

Keywords:

"Immunoassay"[Mesh] OR “antigen test*”[tiab] OR “rapid antigen detection test*”[tiab] OR “antigen detection”[tiab] OR “rapid antigen”[tiab] OR “rapid test*”[tiab] OR “Antigen rapid test*”[tiab] OR “ELISA Test*”[tiab] OR “rapid detection test*”[tiab] OR “[rapid diagnostic test](https://en.wikipedia.org/wiki/Rapid_diagnostic_test)*”[tiab] OR “rapid diagnosis”[tiab]

MeSH: "Immunoassay"[Mesh]

Concept 3: RT-PCR

Keywords:

"Reverse Transcriptase Polymerase Chain Reaction"[Mesh] OR “Reverse Transcriptase PCR” [tiab] OR RT-PCR [tiab] OR PCR[tiab] OR qPCR[tiab] OR qRT-PCR[tiab]

MeSH: "Reverse Transcriptase Polymerase Chain Reaction"[Mesh]

**Characteristics of studies:**

Most of the studies (n=27) used nasopharyngeal swabs, others (n=8) used nasal swabs and the rest (n=4) used either naso-oropharyngeal or nasopharyngeal and throat swabs.

All Included studies are diagnostic studies that assess Ag-RDT products for the detection of COVID-19 nucleocapsid proteins, therefore all of them were published in 2020 and 2021 after the pandemic has emerged.

**Grouping:**

Some approximations were made and the closest range to the defined groups was taken. In case a study provided two datasets for an identical population for the same group, we included the closest to our selected group threshold (e.g., given 30 Ct and 35 Ct for the same population and the same RDT test, only 30 Ct was included to group five).

**Low data in group 1:**

We believe that studies might have not stratified at lower values (Ct <20) because they had reached high sensitivities (>95%) at higher thresholds. Therefore, there was not a reason to consider stratifying at lower thresholds (e.g., Ct <20). This might be a valid explanation for not finding enough number of studies with a Ct value falling under group one category. An example supporting our theory would be Pilarowski and colleagues (2020) findings.^1^ In this study, 100% sensitivity was reached at Ct <30, meaning stratification at a Ct <25 or Ct <20 thresholds would not provide any higher sensitivity.

**Tables**

**Supplementary Table 1 – Extracted datasets from the included studies and their QUADAS-2 scores**

| **Study** | **Ct Group** | **Ag-RDT(+)**  **RT-PCR(+)** | **Ag-RDT(+)**  **RT-PCR(-)** | **Ag-RDT(-)**  **RT-PCR(-)** | **Ag-RDT(-)**  **RT-PCR(+)** | **PPA (%)** | **NPA (%)** | **QUADAS-2 score** | **Value** |
| --- | --- | --- | --- | --- | --- | --- | --- | --- | --- |
| Pena-Rodríguez et al (2021)^2^ | 5 | 79 | 0 | 265 | 25 | 76 | 100 | 11 | 1 |
| Pilarowski et al (2021)^3^ | 5 | 14 | 0 | 855 | 1 | 93·3 | 100 | 9 | 1 |
| Pollock et al (2021)^4^ | 5 | 205 | 12 | 2004 | 9 | 95·8 | 99·4 | 10 | 1 |
| Pollock et al (2021)^4^ | 4 | 149 | 12 | 2004 | 1 | 99·3 | 99·4 | 10 | 1 |
| Pilarowski et al (2020)^1^ | 5 | 171 | 3 | 3062 | 0 | 100 | 99·9 | 9 | 1 |
| Albert et al (2021)^5^ | 5 | 43 | 0 | 358 | 11 | 79·6 | 100 | 11 | 1 |
| Aoki et al (December 23, 2020)^6^ | 5 | 25 | 2 | 64 | 38 | 39·7 | 97 | 6 | 0 |
| Aoki et al (December 3, 2020)^7^ | 5 | 22 | 8 | 516 | 2 | 91·7 | 98·5 | 6 | 0 |
| Bulilete et al (2021)^8^ | 5 | 96 | 2 | 1227 | 29 | 76·8 | 99·8 | 12 | 1 |
| Bulilete et al (2021)^8^ | 4 | 88 | 2 | 1227 | 13 | 87·1 | 99·8 | 12 | 1 |
| Ristic et al (2021)^9^ | 5 | 25 | 0 | 77 | 18 | 58·1 | 100 | 11 | 1 |
| Möckel et al (2021)a^10^ | 5 | 67 | 0 | 182 | 22 | 75·3 | 100 | 11 | 1 |
| Möckel et al (2021)b^10^ | 5 | 18 | 1 | 176 | 7 | 72 | 99·4 | 11 | 1 |
| Merino et al (2021)^11^ | 4 | 184 | 7 | 592 | 1 | 99·5 | 98·8 | 10 | 0 |
| Merino et al (2021)^11^ | 5 | 325 | 7 | 592 | 34 | 90·5 | 98.8 | 10 | 0 |
| Nalumansi et al (2021)^12^ | 5 | 34 | 13 | 159 | 3 | 91·9 | 92·4 | 9 | 1 |
| Nalumansi et al (2021)^12^ | 3 | 29 | 13 | 159 | 24 | 54·7 | 92·4 | 9 | 1 |
| Osterman et al (2021)^13^ | 5 | 224 | 9 | 377 | 221 | 50·3 | 97·7 | 10 | 1 |
| Osterman et al (2021)^13^ | 5 | 173 | 8 | 352 | 208 | 45·4 | 97·8 | 10 | 1 |
| Okoye et al (2021)^14^ | 4 | 23 | 0 | 2593 | 1 | 95·8 | 100 | 10 | 1 |
| Okoye et al (2021)^14^ | 5 | 24 | 0 | 2593 | 12 | 66·7 | 100 | 10 | 1 |
| Okoye et al (2021)^14^ | 3 | 0 | 0 | 2593 | 9 | 0 | 100 | 10 | 1 |
| Young et al (2020)^15^ | 5 | 29 | 1 | 212 | 9 | 76·3 | 99·5 | 11 | 0 |
| Villaverde et al (2021)^16^ | 5 | 35 | 3 | 1540 | 42 | 45·5 | 99·8 | 9 | 0 |
| Scohy et al (2021)^17^ | 4 | 10 | 0 | 42 | 0 | 100 | 100 | 10 | 1 |
| Scohy et al (2021)^17^ | 5 | 24 | 0 | 42 | 10 | 70·6 | 100 | 10 | 1 |
| Porte et al (2021)^18^ | 4 | 27 | 1 | 31 | 0 | 100 | 96·9 | 11 | 1 |
| Porte et al (2021)^18^ | 5 | 30 | 1 | 31 | 2 | 93·8 | 96·9 | 11 | 1 |
| Porte et al (2021)^18^ | 4 | 27 | 1 | 31 | 0 | 100 | 96·9 | 11 | 1 |
| Porte et al (2021)^18^ | 5 | 29 | 1 | 31 | 3 | 90·6 | 96·9 | 11 | 1 |
| Porte et al (2020)^19^ | 4 | 52 | 0 | 45 | 0 | 100 | 100 | 11 | 1 |
| Porte et al (2020)^19^ | 5 | 77 | 0 | 45 | 5 | 93·9 | 100 | 11 | 1 |
| Pray et al (2021)^20^ | 5 | 39 | 16 | 1025 | 18 | 68·4 | 98·5 | 8 | 0 |
| Pérez et al (2021)^21^ | 4 | 79 | 0 | 150 | 5 | 94 | 100 | 10 | 0 |
| Pérez et al (2021)^21^ | 5 | 91 | 0 | 150 | 79 | 53·5 | 100 | 10 | 0 |
| Pérez et al (2021)^21^ | 4 | 81 | 0 | 150 | 3 | 96·4 | 100 | 10 | 0 |
| Pérez et al (2021)^21^ | 5 | 102 | 0 | 150 | 68 | 60 | 100 | 10 | 0 |
| Prince et al (2021)^22^ | 5 | 157 | 4 | 3116 | 142 | 52·5 | 99·9 | 10 | 1 |
| Jääskeläinen et al (2021)^23^ | 4 | 88 | 0 | 40 | 1 | 98·9 | 100 | 9 | 0 |
| Jääskeläinen et al (2021)^23^ | 2 | 28 | 0 | 40 | 6 | 82·4 | 100 | 9 | 0 |
| Jääskeläinen et al (2021)^23^ | 5 | 116 | 0 | 40 | 7 | 94·3 | 100 | 9 | 0 |
| Jääskeläinen et al (2021)^23^ | 3 | 3 | 0 | 40 | 22 | 12 | 100 | 9 | 0 |
| Jääskeläinen et al (2021)^23^ | 4 | 96 | 0 | 40 | 1 | 99 | 100 | 9 | 0 |
| Jääskeläinen et al (2021)^23^ | 2 | 24 | 0 | 40 | 11 | 68·6 | 100 | 9 | 0 |
| Jääskeläinen et al (2021)^23^ | 5 | 120 | 0 | 40 | 12 | 90·9 | 100 | 9 | 0 |
| Jääskeläinen et al (2021)^23^ | 3 | 8 | 0 | 40 | 18 | 30·8 | 100 | 9 | 0 |
| Jääskeläinen et al (2021)^23^ | 4 | 90 | 0 | 38 | 2 | 97·8 | 100 | 9 | 0 |
| Jääskeläinen et al (2021)^23^ | 2 | 26 | 0 | 38 | 8 | 76·5 | 100 | 9 | 0 |
| Jääskeläinen et al (2021)^23^ | 5 | 116 | 0 | 38 | 10 | 92·1 | 100 | 9 | 0 |
| Jääskeläinen et al (2021)^23^ | 3 | 10 | 0 | 38 | 16 | 38·5 | 100 | 9 | 0 |
| Kobayashi et al (2021)^24^ | 5 | 56 | 8 | 192 | 18 | 75·7 | 96 | 11 | 1 |
| Kohmer et al (2021)^25^ | 4 | 15 | 1 | 25 | 1 | 93·8 | 96·2 | 9 | 0 |
| Kohmer et al (2021)^25^ | 2 | 11 | 1 | 25 | 12 | 47·8 | 96·2 | 9 | 0 |
| Kohmer et al (2021)^25^ | 5 | 26 | 1 | 25 | 13 | 66·7 | 96·2 | 9 | 0 |
| Kohmer et al (2021)^25^ | 3 | 3 | 1 | 25 | 32 | 8·6 | 96·2 | 9 | 0 |
| Kohmer et al (2021)^25^ | 4 | 16 | 0 | 26 | 0 | 100 | 100 | 9 | 0 |
| Kohmer et al (2021)^25^ | 2 | 14 | 0 | 26 | 9 | 60·9 | 100 | 9 | 0 |
| Kohmer et al (2021)^25^ | 5 | 30 | 0 | 26 | 9 | 76·9 | 100 | 9 | 0 |
| Kohmer et al (2021)^25^ | 3 | 2 | 0 | 26 | 33 | 5·7 | 100 | 9 | 0 |
| Kohmer et al (2021)^25^ | 4 | 14 | 0 | 26 | 2 | 87·5 | 100 | 9 | 0 |
| Kohmer et al (2021)^25^ | 2 | 3 | 0 | 26 | 20 | 13 | 100 | 9 | 0 |
| Kohmer et al (2021)^25^ | 5 | 17 | 0 | 26 | 22 | 43·6 | 100 | 9 | 0 |
| Kohmer et al (2021)^25^ | 3 | 1 | 0 | 26 | 34 | 2·9 | 100 | 9 | 0 |
| Kohmer et al (2021)^25^ | 4 | 16 | 0 | 26 | 0 | 100 | 100 | 9 | 0 |
| Kohmer et al (2021)^25^ | 2 | 17 | 0 | 26 | 6 | 73·9 | 100 | 9 | 0 |
| Kohmer et al (2021)^25^ | 5 | 33 | 0 | 26 | 6 | 84·6 | 100 | 9 | 0 |
| Krüttgen et al (2021)^26^ | 3 | 4 | 0 | 26 | 31 | 11·4 | 100 | 9 | 0 |
| Krüttgen et al (2021)^26^ | 4 | 17 | 3 | 72 | 0 | 100 | 96 | 10 | 0 |
| Krüttgen et al (2021)^26^ | 2 | 19 | 3 | 72 | 1 | 95 | 96 | 10 | 0 |
| Krüttgen et al (2021)^26^ | 3 | 15 | 3 | 72 | 23 | 39·5 | 96 | 10 | 0 |
| Linares et al (2020) ^27^ | 5 | 44 | 0 | 195 | 16 | 73·3 | 100 | 10 | 0 |
| Chaimayo et al (2020)^28^ | 5 | 59 | 5 | 389 | 1 | 98·3 | 98·7 | 10 | 1 |
| Courtellemont et al (2021)^29^ | 5 | 65 | 0 | 127 | 1 | 98·5 | 100 | 10 | 1 |
| Courtellemont et al (2021)^29^ | 3 | 117 | 0 | 127 | 4 | 96·7 | 100 | 10 | 1 |
| Drain et al (2021)^30^ | 5 | 39 | 5 | 210 | 1 | 97·5 | 97·7 | 8 | 0 |
| Favresse et al (2021)^31^ | 1 | 35 | 4 | 104 | 1 | 97·2 | 96·3 | 12 | 1 |
| Favresse et al (2021)^31^ | 4 | 54 | 4 | 104 | 4 | 93·1 | 96·3 | 12 | 1 |
| Favresse et al (2021)^31^ | 2 | 25 | 4 | 104 | 16 | 61 | 96·3 | 12 | 1 |
| Favresse et al (2021)^31^ | 5 | 60 | 4 | 104 | 17 | 77·9 | 96·3 | 12 | 1 |
| Favresse et al (2021)^31^ | 3 | 1 | 4 | 104 | 2 | 33·3 | 96·3 | 12 | 1 |
| Favresse et al (2021)^31^ | 1 | 33 | 1 | 107 | 3 | 91·7 | 99·1 | 12 | 1 |
| Favresse et al (2021)^31^ | 4 | 54 | 1 | 107 | 4 | 93.1 | 99·1 | 12 | 1 |
| Favresse et al (2021)^31^ | 2 | 31 | 1 | 107 | 10 | 75·6 | 99·1 | 12 | 1 |
| Favresse et al (2021)^31^ | 5 | 64 | 1 | 107 | 13 | 83·1 | 99·1 | 12 | 1 |
| Favresse et al (2021)^31^ | 3 | 0 | 1 | 107 | 3 | 0 | 99·1 | 12 | 1 |
| Favresse et al (2021)^31^ | 1 | 35 | 6 | 102 | 1 | 97·2 | 94·4 | 12 | 1 |
| Favresse et al (2021)^31^ | 4 | 56 | 6 | 102 | 2 | 96·6 | 94·4 | 12 | 1 |
| Favresse et al (2021)^31^ | 2 | 34 | 6 | 102 | 7 | 82·9 | 94·4 | 12 | 1 |
| Favresse et al (2021)^31^ | 5 | 69 | 6 | 102 | 8 | 89·6 | 94·4 | 12 | 1 |
| Favresse et al (2021)^31^ | 3 | 2 | 6 | 102 | 1 | 66·7 | 94·4 | 12 | 1 |
| Favresse et al (2021)^31^ | 1 | 35 | 1 | 107 | 1 | 97·2 | 99·1 | 12 | 1 |
| Favresse et al (2021)^31^ | 4 | 56 | 1 | 107 | 2 | 96·6 | 99·1 | 12 | 1 |
| Favresse et al (2021)^31^ | 2 | 31 | 1 | 107 | 10 | 75·6 | 99·1 | 12 | 1 |
| Favresse et al (2021)^31^ | 5 | 66 | 1 | 107 | 11 | 85·7 | 99·1 | 12 | 1 |
| Favresse et al (2021)^31^ | 3 | 0 | 1 | 107 | 3 | 0 | 99·1 | 12 | 1 |
| Fenollar et al (2021)^32^ | 1 | 58 | 7 | 130 | 1 | 98·3 | 94·9 | 10 | 0 |
| Fenollar et al (2021)^32^ | 4 | 107 | 7 | 130 | 4 | 96·4 | 94·9 | 10 | 0 |
| Fenollar et al (2021)^32^ | 2 | 88 | 7 | 130 | 17 | 83·8 | 94·9 | 10 | 0 |
| Fenollar et al (2021)^32^ | 5 | 146 | 7 | 130 | 18 | 89 | 94·9 | 10 | 0 |
| Fenollar et al (2021)^32^ | 3 | 8 | 7 | 130 | 32 | 20 | 94·9 | 10 | 0 |
| Gili et al (2021)a^33^ | 5 | 86 | 11 | 120 | 9 | 90·5 | 91·6 | 9 | 1 |
| Gili et al (2021)b^33^ | 5 | 90 | 130 | 1518 | 0 | 100 | 92·1 | 9 | 1 |
| Ishii et al (2021)a^34^ | 5 | 22 | 1 | 460 | 2 | 91·7 | 99·8 | 10 | 1 |
| Ishii et al (2021)b^34^ | 5 | 10 | 0 | 260 | 1 | 90·9 | 100 | 10 | 1 |
| Gremmels et al (2021)a^35^ | 5 | 101 | 0 | 1228 | 38 | 72·7 | 100 | 12 | 1 |
| Gremmels et al (2021)b^35^ | 5 | 51 | 0 | 145 | 12 | 81 | 100 | 12 | 1 |
| Gupta et al (2020)^36^ | 5 | 63 | 1 | 252 | 14 | 81·8 | 99·6 | 11 | 0 |
| Ciotti et al (2021)^37^ | 5 | 12 | 0 | 11 | 27 | 30·7 | 100 | 11 | 1 |
| Hirotsu et al (2021)^38^ | 5 | 37 | 0 | 989 | 3 | 92·5 | 100 | 8 | 1 |
| Yamamoto et al (2020)^39^ | 5 | 52 | 1 | 100 | 76 | 41 | 99 | 6 | 0 |

Ag-RDT(-)RT-PCR(-) and Ag-RDT(+)RT-PCR(-) values are only considered for the highest Ct in the studies. Value=1 if RT-PCR detection threshold is reported in the paper. Value=0 if RT-PCR detection threshold is not reported in the paper. Group 1= Ct <20. Group 2= Ct 20-30. Group 3= Ct >30. Group 4= Ct <25. Group 5= Ct <30. Full datasets are available with the corresponding author on reasonable request.

**Supplementary table 2: Sample size and number of datasets per Ct group and product**

| Ct group | Product | Datasets | N |
| --- | --- | --- | --- |
| 1 | Total | 5 | 772 |
| 2 | Panbio | 3 | 463 |
| 2 | Roche | 3 | 293 |
| 2 | Other | 4 | 396 |
| 2 | Total | 13 | 1350 |
| 3 | Panbio | 3 | 352 |
| 3 | Roche | 3 | 285 |
| 3 | Other | 5 | 592 |
| 3 | Total | 15 | 1646 |
| 4 | Panbio | 6 | 2892 |
| 4 | Roche | 3 | 300 |
| 4 | Other | 8 | 858 |
| 4 | Total | 23 | 9200 |
| 5 | BinaxNOW | 5 | 12384 |
| 5 | Panbio | 11 | 7144 |
| 5 | Roche | 5 | 1554 |
| 5 | Sofia | 3 | 1325 |
| 5 | Lumipulse | 6 | 4300 |
| 5 | StandardQ | 6 | 1654 |
| 5 | Other | 15 | 2951 |
| 5 | Total | 53 | 21632 |

N=number of samples. Group 1= Ct <20. Group 2= Ct 20-30. Group 3= Ct >30. Group 4= Ct <25. Group 5= Ct <30

**Supplementary table 3: List of product groups and manufacturing companies**

| Product groups | Company/Description |
| --- | --- |
| BinaxNOW | Abbott |
| Panbio | Abbott |
| Roche | Roche Diagnostics |
| Sofia | Quidel |
| StandardQ | SD Biosensor |
| Lumipulse | Fujirebio |
| Other | All other products |
| Total | Total of all groups |

**Doi Plots**

**Supplementary figure 1: Doi plot for PPA of Group 1 Ct <20**

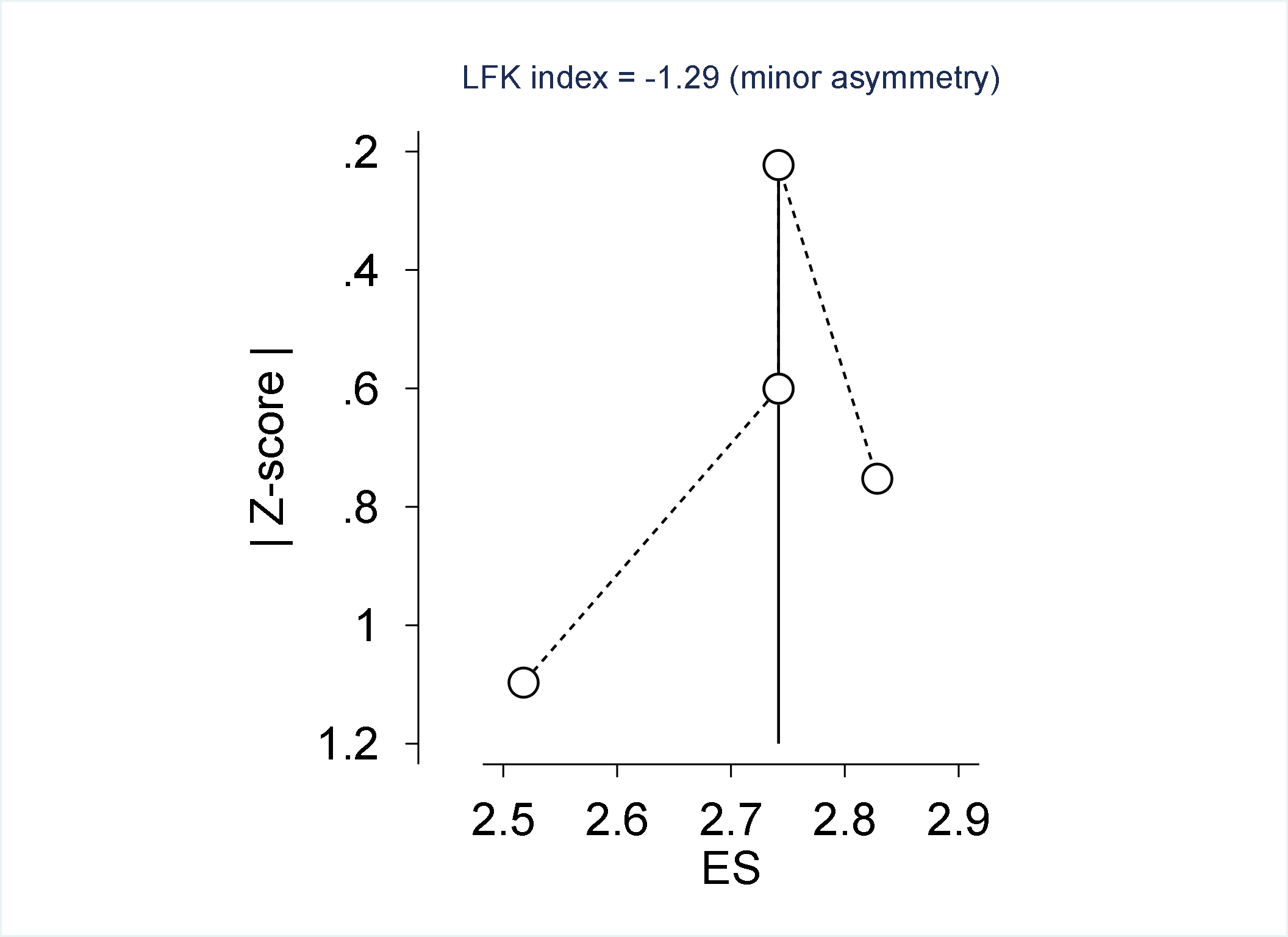

**Supplementary figure 2: Doi plot for PPA of Group 2 Ct 20-30**

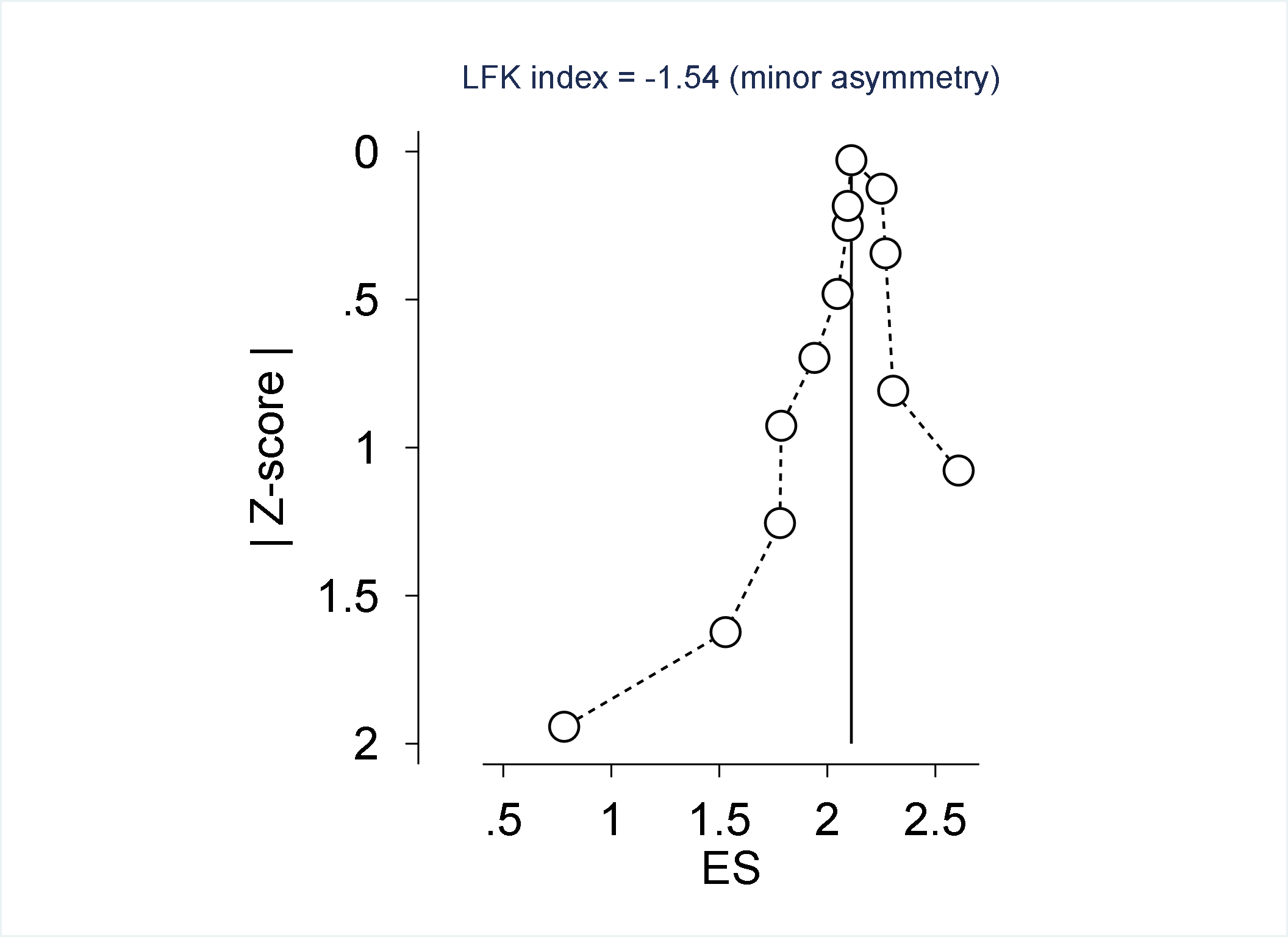

**Supplementary figure 3: Doi plot for PPA of Group 3 Ct >30**

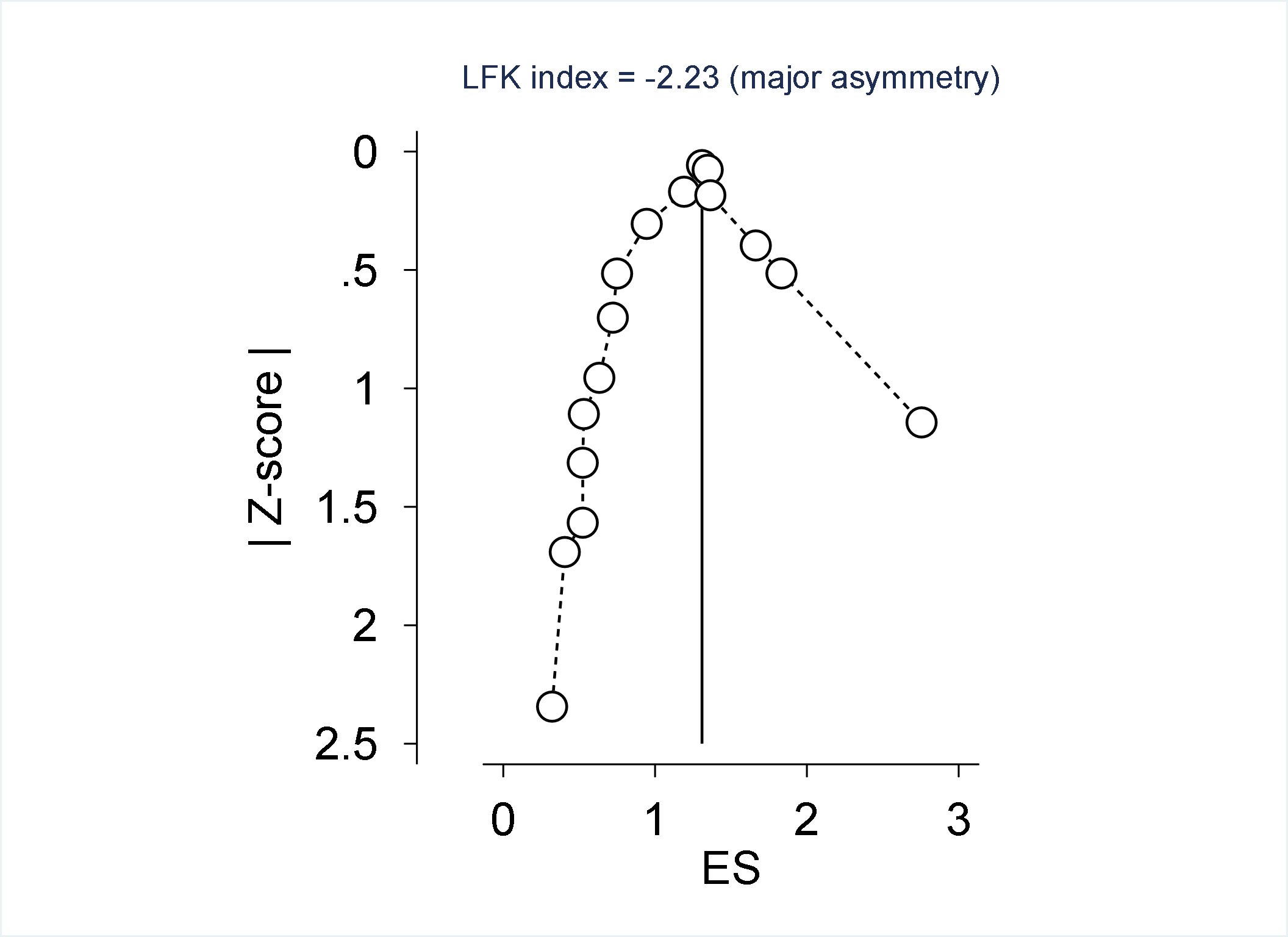

**Supplementary figure 4: Doi plot for PPA of Group 4 Ct <25**

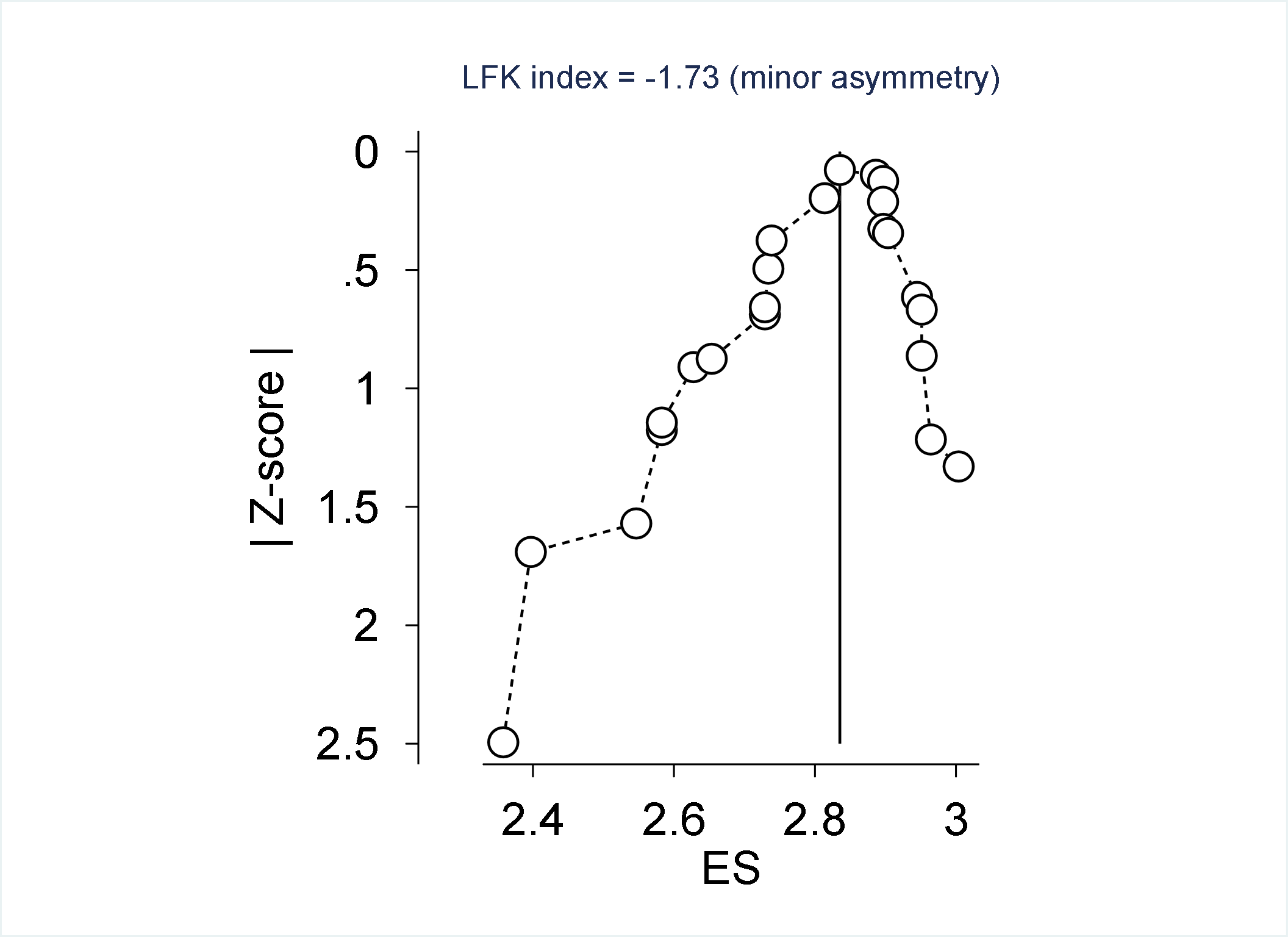

**Supplementary figure 5: Doi plot for PPA of Group 5 Ct <30**

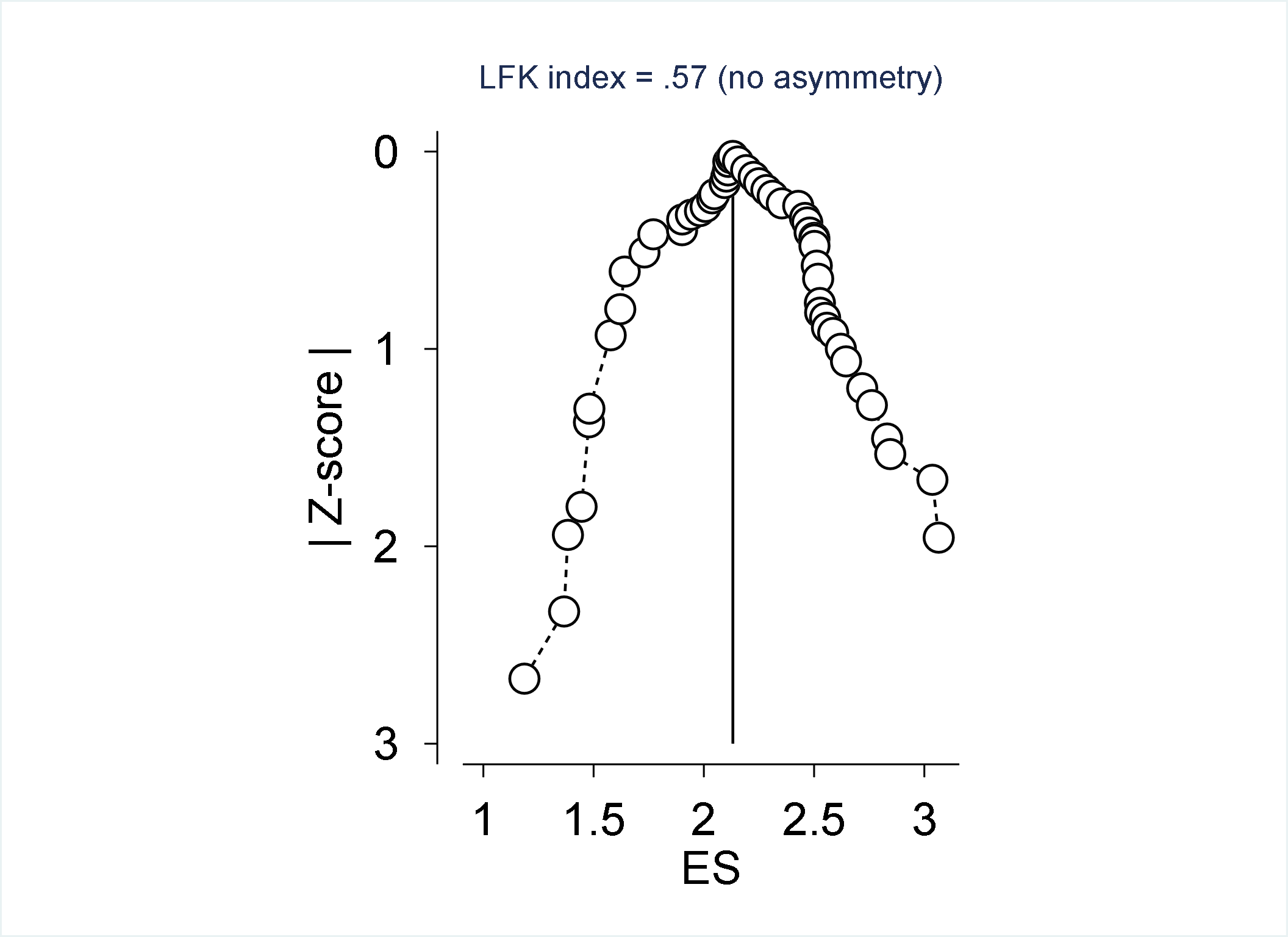

**Supplementary figure 6: Doi plot for NPA of all Products**

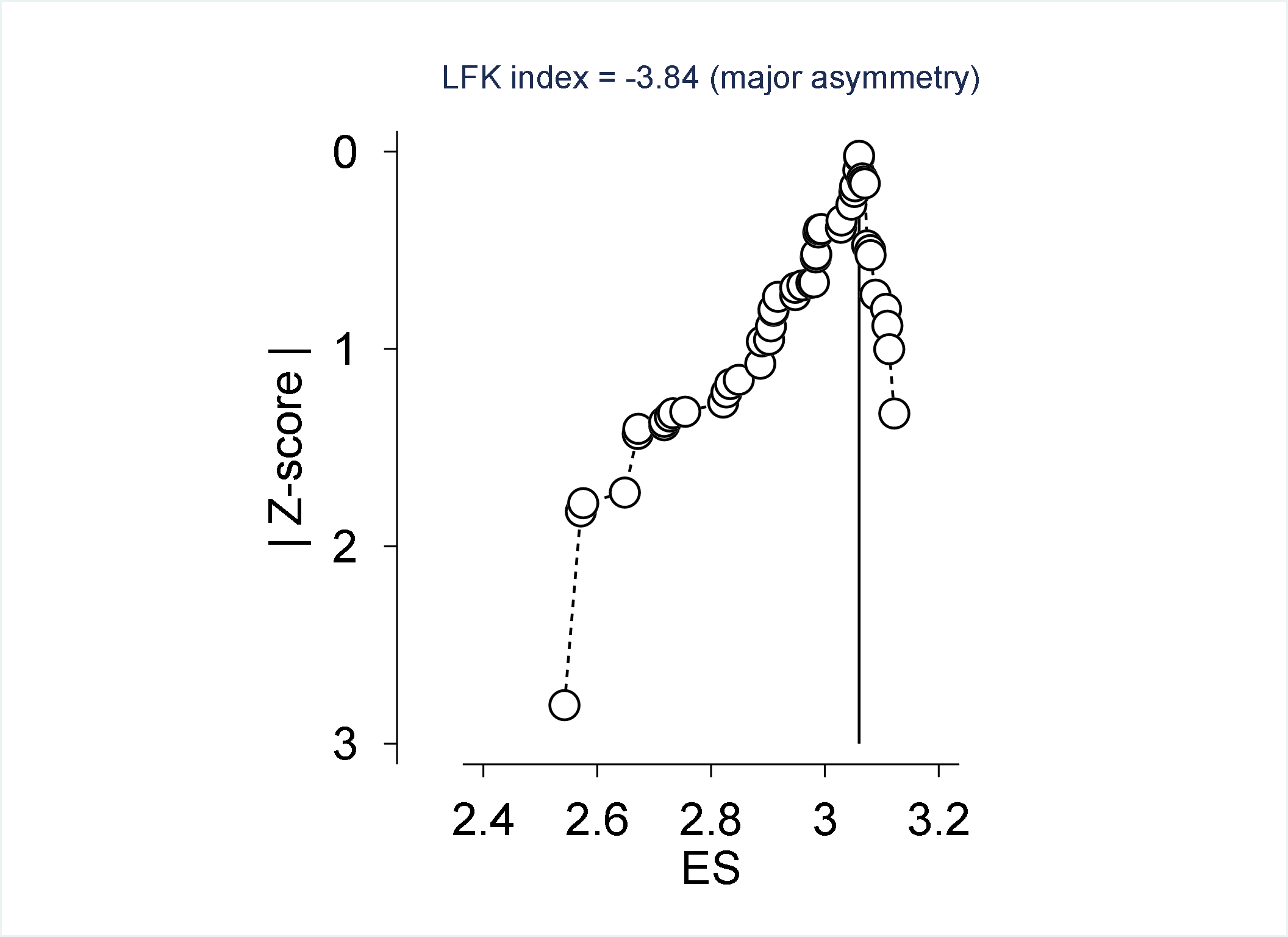
